# Exploring alternative medicine options for the prevention or treatment of coronavirus disease 2019 (COVID-19)- A systematic scoping review

**DOI:** 10.1101/2020.05.14.20101352

**Authors:** Amrita Nandan, Santosh Tiwari, Vishwas Sharma

## Abstract

**Background:** Coronavirus disease 2019 (COVID-19) is caused by coronavirus 2 (SARS-CoV-2). Symptoms include fever, cough, shortness of breath, muscle pain, pneumonia, and multiorgan failure. The infection spreads from one person to another via respiratory droplets. Alternative medicine (AMs) viz., Ayurveda, Homeopathy, Unani, and Traditional Chinese Medicine (TCM), are being promoted for the prevention of COVID-19. The aim of this systematic scoping review was to identify and summarize the scientific evidences promoting the use of AMs for the prevention of COVID-19.

**Methods:** A comprehensive search of electronic search engines (PubMed and Web of Science) was performed. In addition, freewheeling searches of the government health ministries and government websites was done to retrieve the available information. Records available until 12^th^ March 2020 were considered. Reports proposing the use of AMs for prevention or treatment of COVID-19 across all countries were included. Screening (primary and secondary) of the records and data extraction from the eligible studies were done by a single reviewer followed by a random quality check (10%) by the second reviewer.

**Results:** Overall, 8 records (7 from China and 1 from India) exploring the use of AMs for the prevention or treatment of COVID-19 were identified. Different medicines were explored by different AM systems.

**Conclusions:** Several AMs options are proposed for the prevention or treatment of COVID-19. However, their efficacy and safety still needs scientific validation through rigorous randomized controlled trials. This review may help inform decisions about the importance of research and development of AMs for COVID-19 prevention and treatment.

## Introduction

Coronavirus disease 2019 (COVID-19) is spreading rapidly throughout the world and is infecting people irrespective of their age, sex and ethnicity. The healthcare systems of several countries are already stretched to their limits. The disease spreads via the respiratory droplets and close contact, and the disinfection of cities and communities is not showing to be effective for the control of the disease.^1^ The World Health Organization (WHO) has declared COVID-19 as a global pandemic which is causing critical challenges for the politicians, public health practitioners, scientists, and the medical community to deal with the situation.^2,3^ There is no specific antiviral treatment available in the modern medicine system to treat COVID-19. Despite concerted efforts, a truly effective vaccine is still not available. Under such conditions, it is imperative that resource-starved nations and communities may think for relatively cheaper options of alternative medicines (AMs) for prophylaxis or treat themselves against this virus. Presently, it is a worldwide emergency and the primary objective of mankind at this moment is to survive. The scientific basis of the AM treatment options can be explored and debated later. Mankind cannot allow thousands or millions to perish before modern medicine invents a cure or prophylaxis. In China itself, the total number of confirmed cases treated by Traditional Chinese Medicine (TCM) has reached 60,107.^4^ Indian government ministry of Ayurveda, Yoga & Naturopathy, Unani, Siddha and Homoeopathy (AYUSH) is also proposing Homeopathy and Ayurveda for prophylaxis and Unani medicines for symptomatic management of COVID-19.^5^

There are two broad clinical systems for preventing or treating the disease i) modern medicine, which is experimental medicine based on modern scientific clinical evidence ii) alternative medicine, which comprises (a) Ayurveda: This system uses plant-based medicines with some animal products as well as added minerals; the evidence is largely clinical. (b) Homeopathy: uses an extremely diluted form of numerous substances. Evidence is largely clinical and partly from randomized controlled trials (RCTs). (c) Unani is a Perso-Arabic system of medicine based originally on the teachings of the Greek physicians Hippocrates and Galen. (d) TCM: based on traditional practices and beliefs from China which mostly include herbal formulations besides acupuncture, cupping therapy, gua sha, massage (tui na), bonesetter (die-da), exercise (qigong), and dietary therapy. Recently, articles exploring the use of AMs for COVID-19 prevention or treatment are available in the electronic search engines such as PubMed, Web of Science, etc. This systematic scoping review aims to summarize available evidences on the AM options for the prevention or treatment of COVID-19. Specifically, we aimed to answer the following questions:

1. Is there any recent evidence available for using AMs for the prevention or treatment of COVID-19; if yes then how they are distributed country-wise?
2. Is there any scientific research-based evidence available for the proposed AMs for COVID-19 prevention or treatment?

## Materials and Methods

### Search criteria

A comprehensive literature search was performed following PRISMA guidelines^6,7^ to screen the records on AMs for COVID-19 (Figure 1 and Additional file S1) using the phrase (ayurveda OR traditional chinese medicine OR homeopathy OR unani OR siddha) AND (COVID-19). Briefly, electronic search engines (PubMed and Web of Science) were screened from the inception database to 12^th^ of March 2020. In addition, freewheeling searches of the health ministry and government websites of different countries was done to retrieve all the relevant data. The records issued by the government authorities reflect its acceptance among the society in that country and hence included. One reviewer (A.N.) screened all titles and abstracts and relevant full-text articles. A second researcher (V.S.) performed a quality check of a random selection of 10% of titles, abstracts, and full-text articles. Discrepancies were resolved; when a consensus was not reached, a third researcher was consulted. Relevant data from all included studies were extracted by a single reviewer, using a pre-defined extraction grid, which was subsequently validated by an independent reviewer. The detailed criteria of screening are mentioned on Additional file S1.

**Figure 1.**
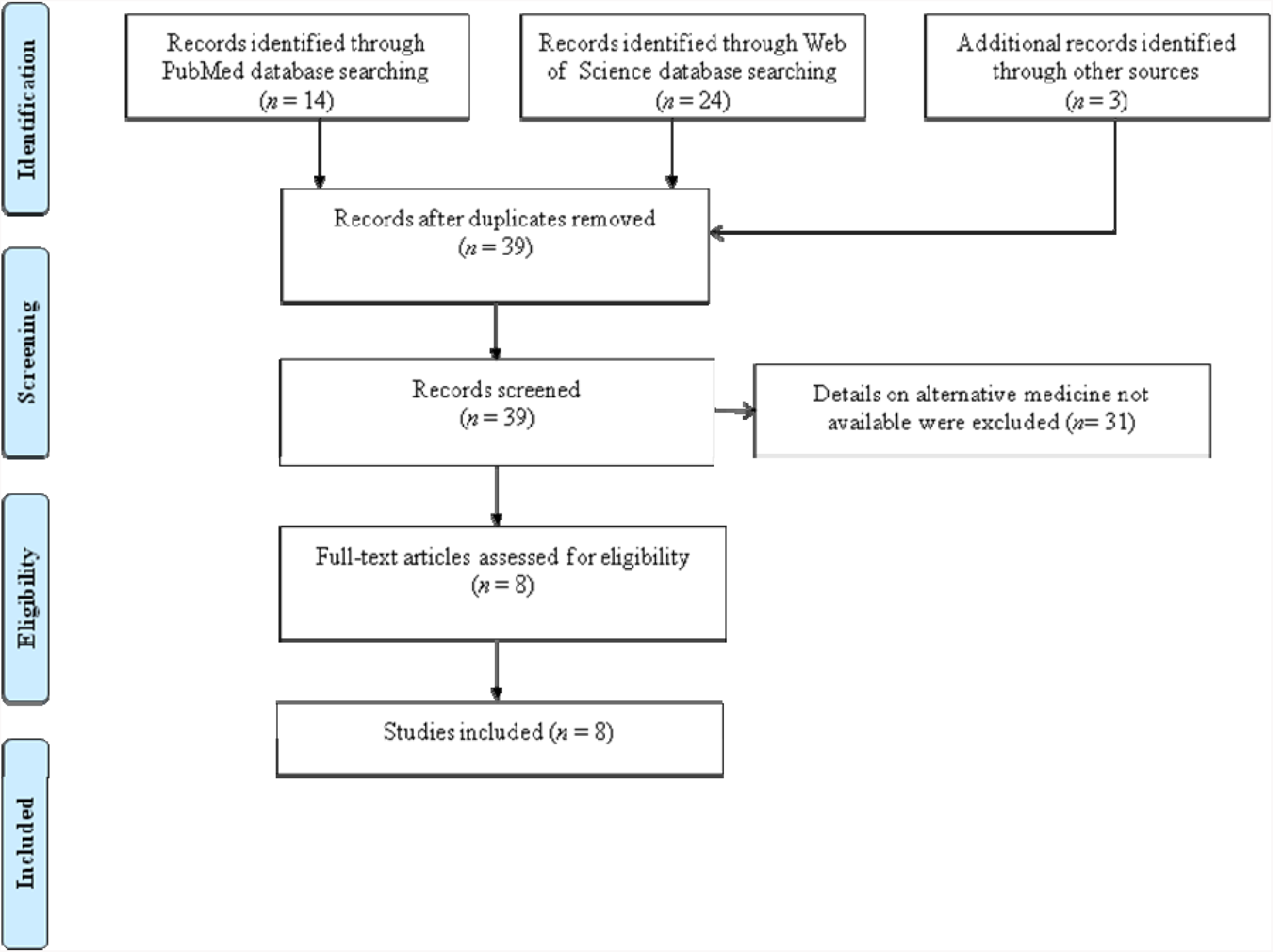
Flow chart describing the included/excluded literature.

The inclusion and exclusion criteria are:

#### Inclusion criteria

i. records where details of AMs (Ayurveda, Homeopathy, Unani, and TCM) on COVID-19 were given, are included for quantitative analysis (Additional file S2).
ii. records whose abstract and full text available were included.
iii. records published in English were included.

#### Exclusion criteria

i. records where details on AMs were not available were excluded.

## Data extraction

From the selected records following were extracted (i) type of AMs system i.e. Ayurveda, Homeopathy, Unani. and TCM (ii) country (iii) clinical treatment (iv) drug dosage recommended(v) drug duration recommended.

## Results

A total of 8 records^4,5,8–13^ were obtained where the promotion of AMs (Ayurveda, Homeopathy, Unani, and TCM) for the prevention or treatment of COVID-19 were described (Additional file S2). Seven records were on TCM and one record covered Ayurveda, Homeopathy, and Unani medicines. Records were either from China (n=7) or India (n=1). Different AMs comprising preventive management and symptomatic management were proposed for the treatment of COVID-19. Diverse drug dose and duration were suggested for specific types of AMs (Table 1). During our search we found only the names of the Ayurvedic, Homeopathic, and Unani Medicines in the record. In order to understand the probable connection of these medicines with COVID-19, we further explored the general properties of promoted AMs whose details are:

**Table 1.**
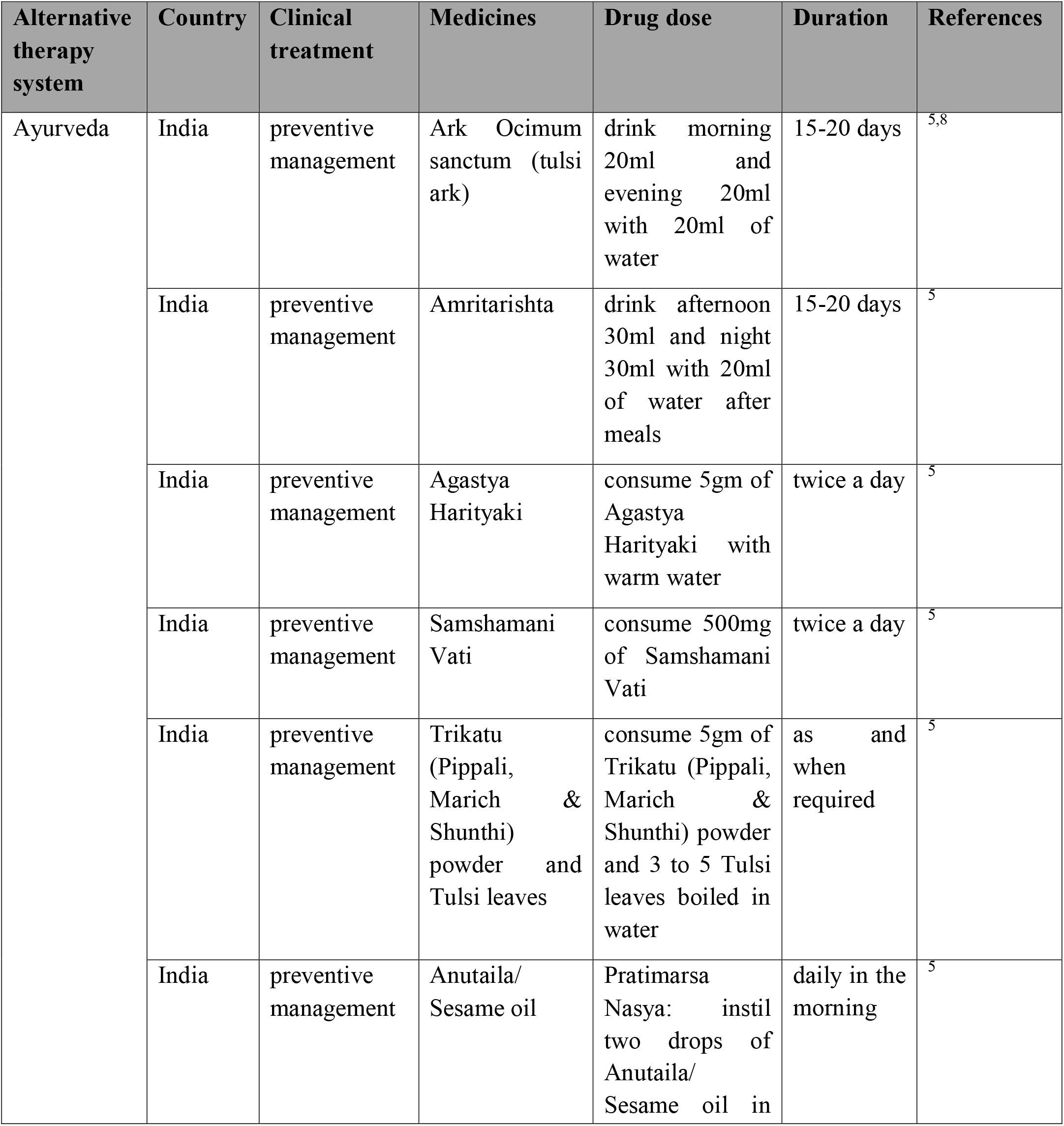
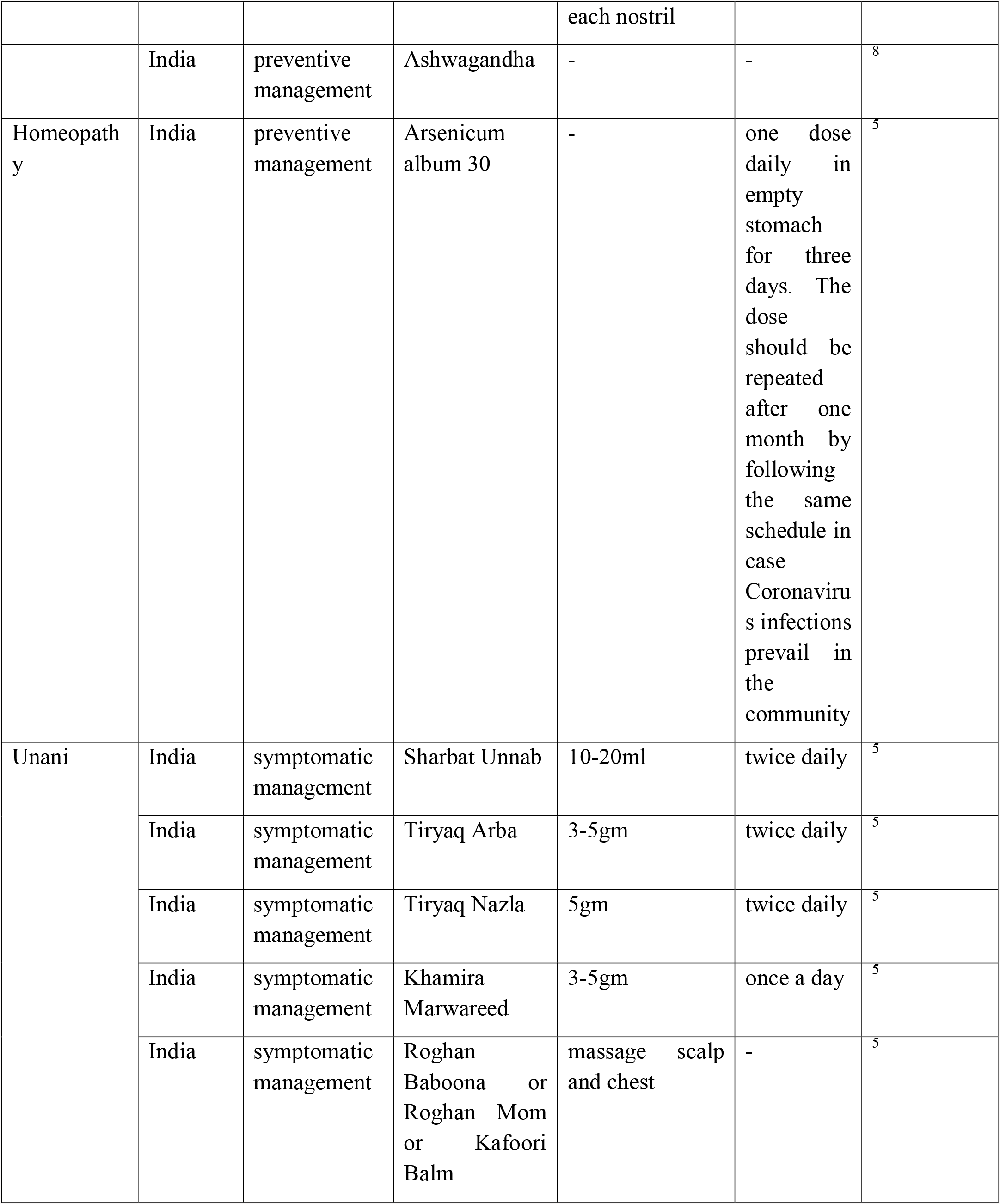
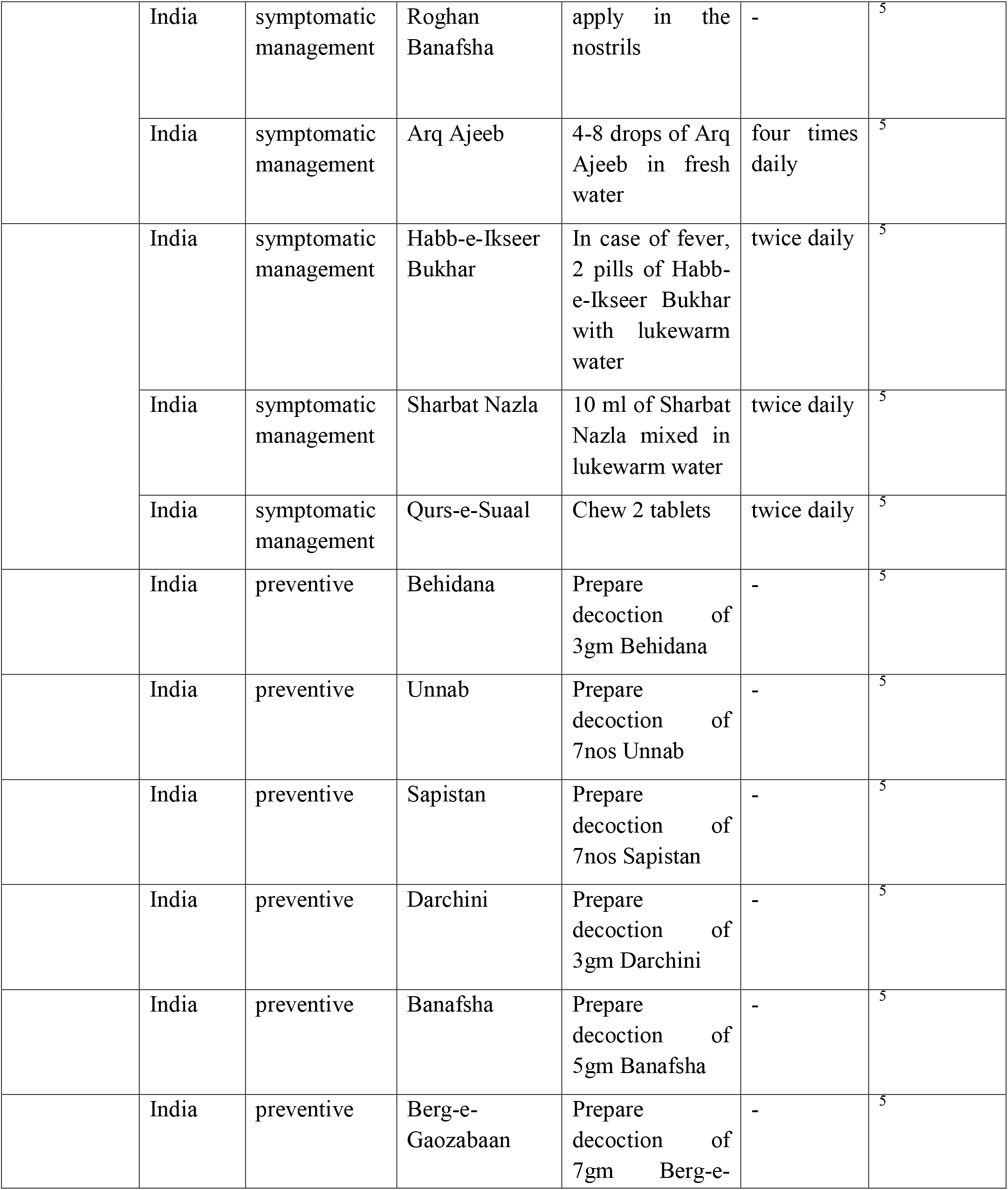
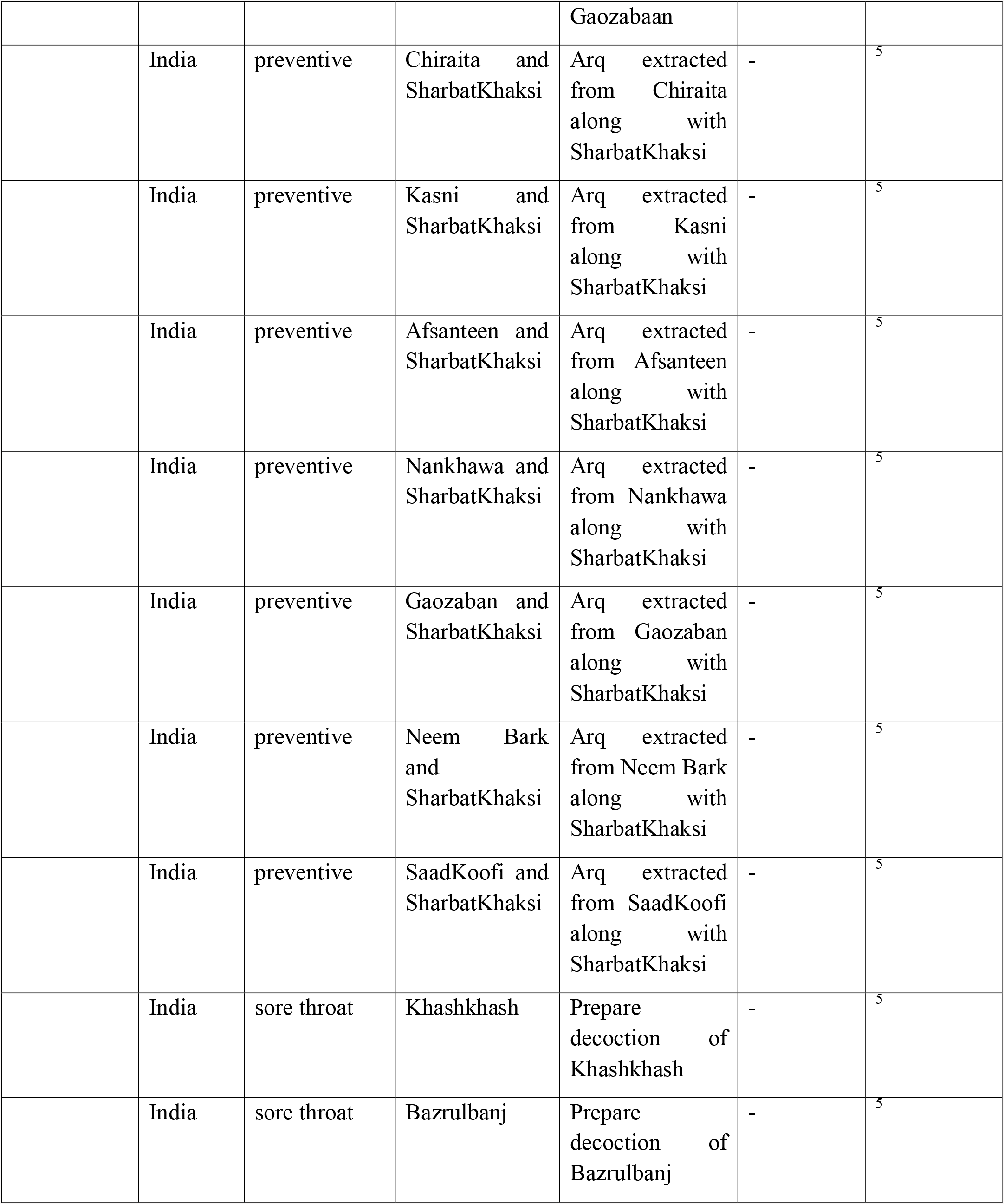
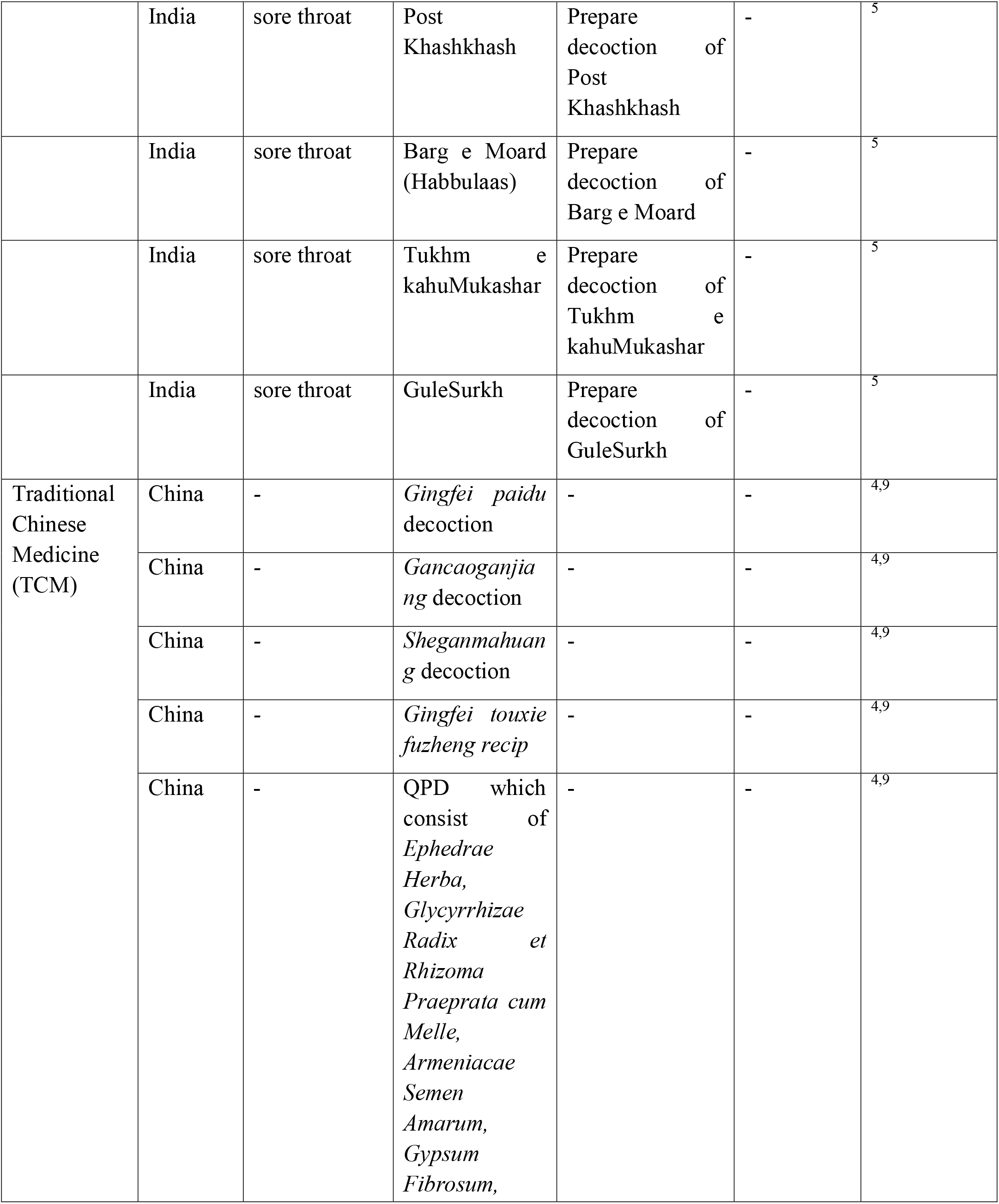
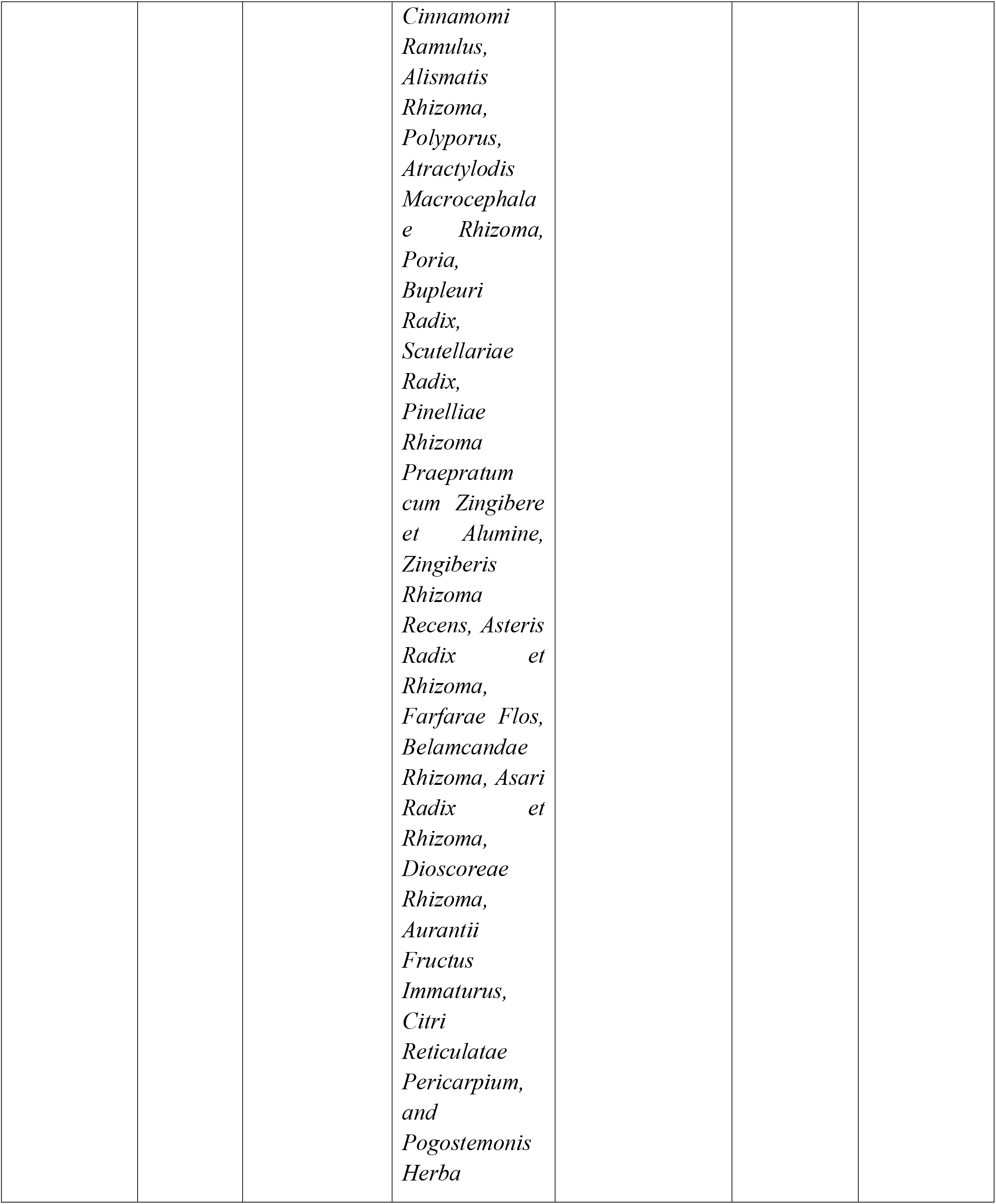
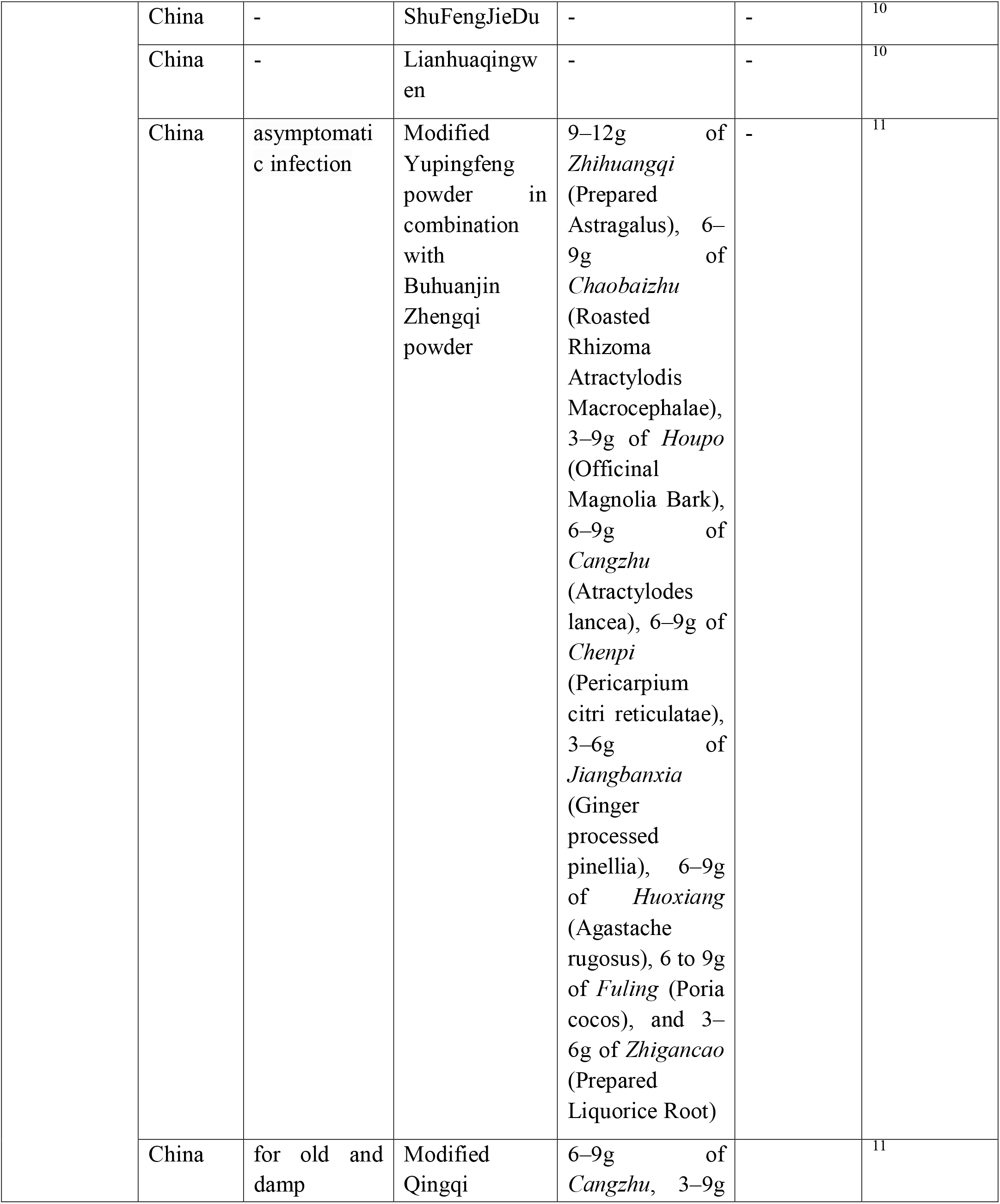
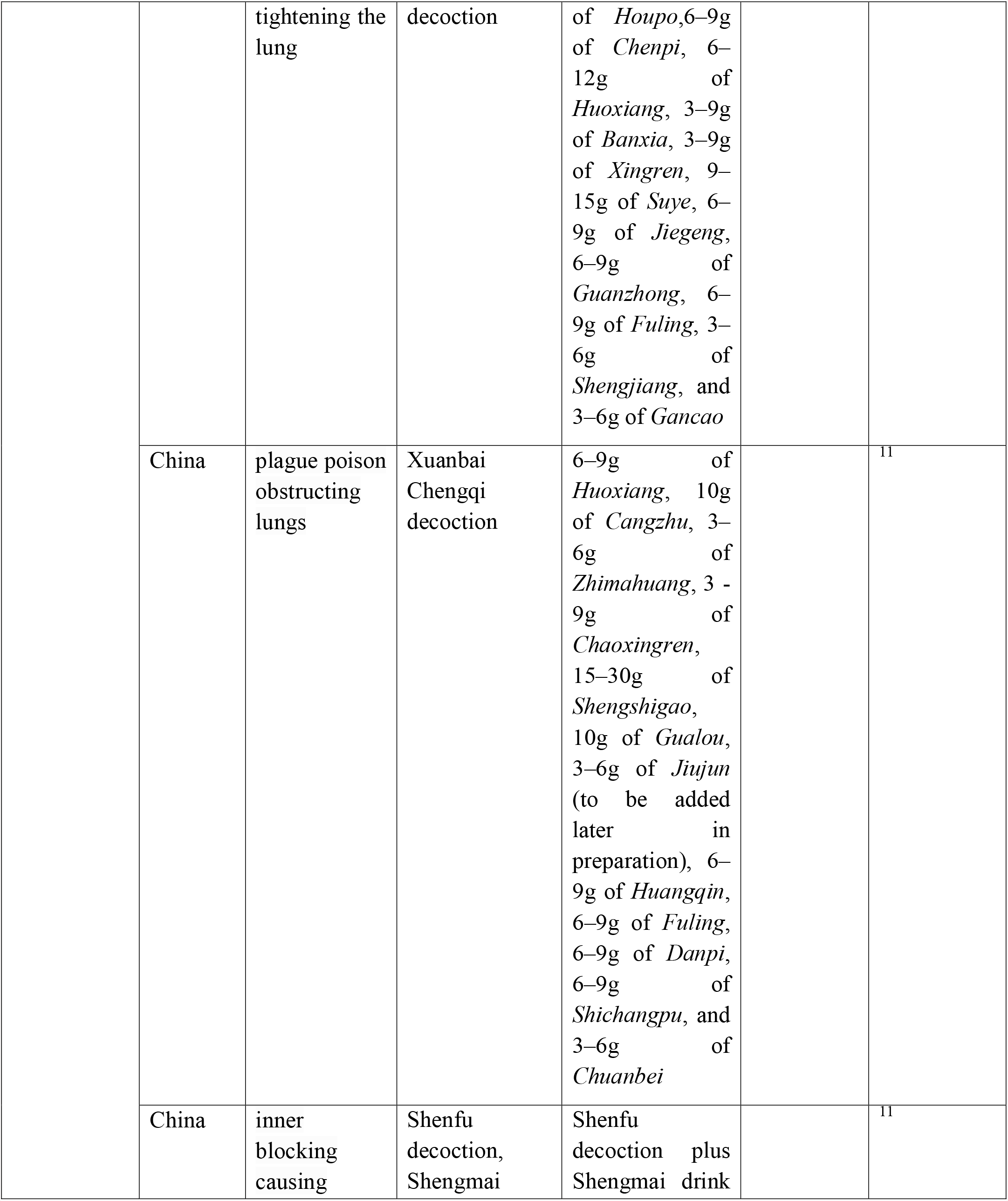
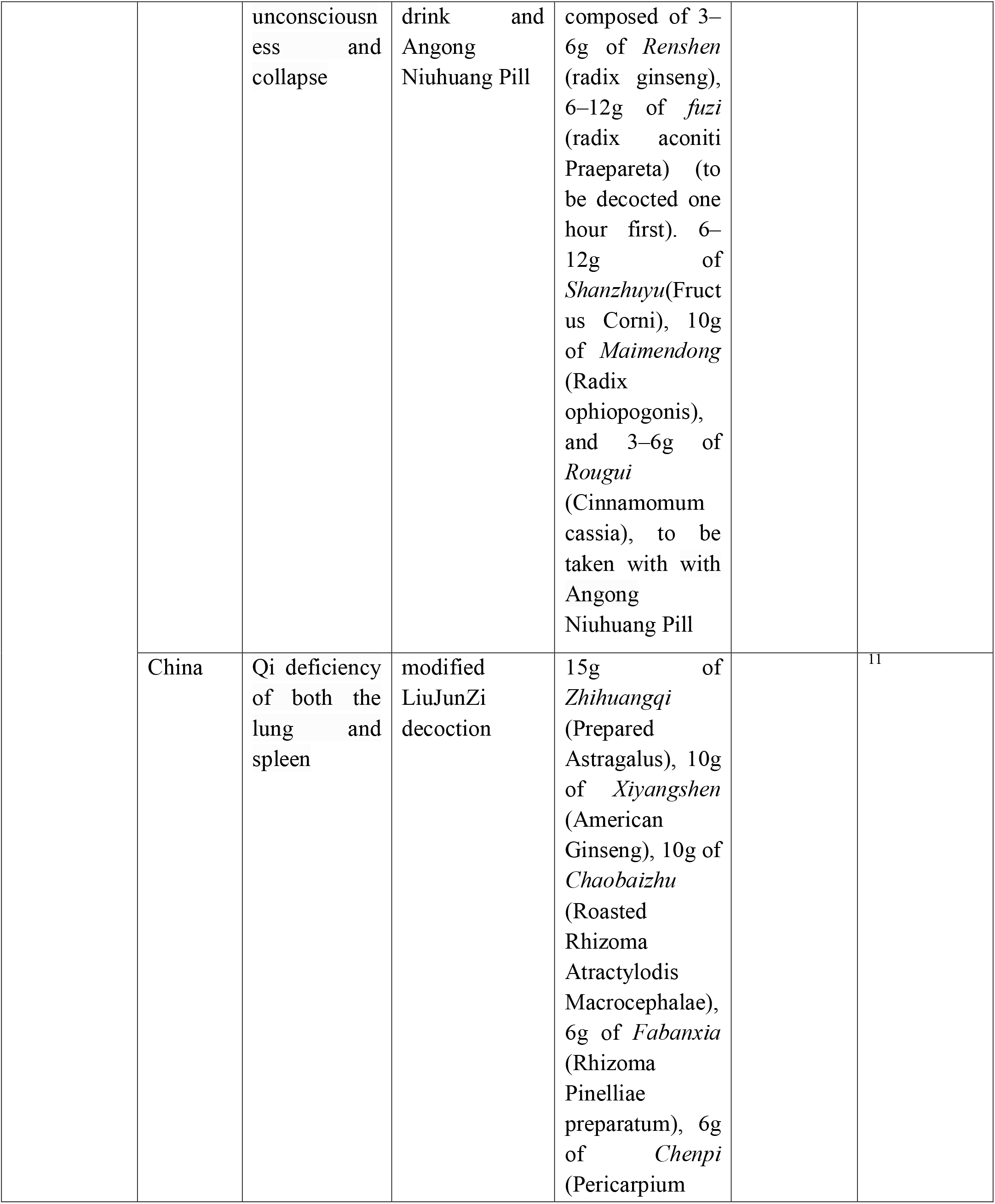
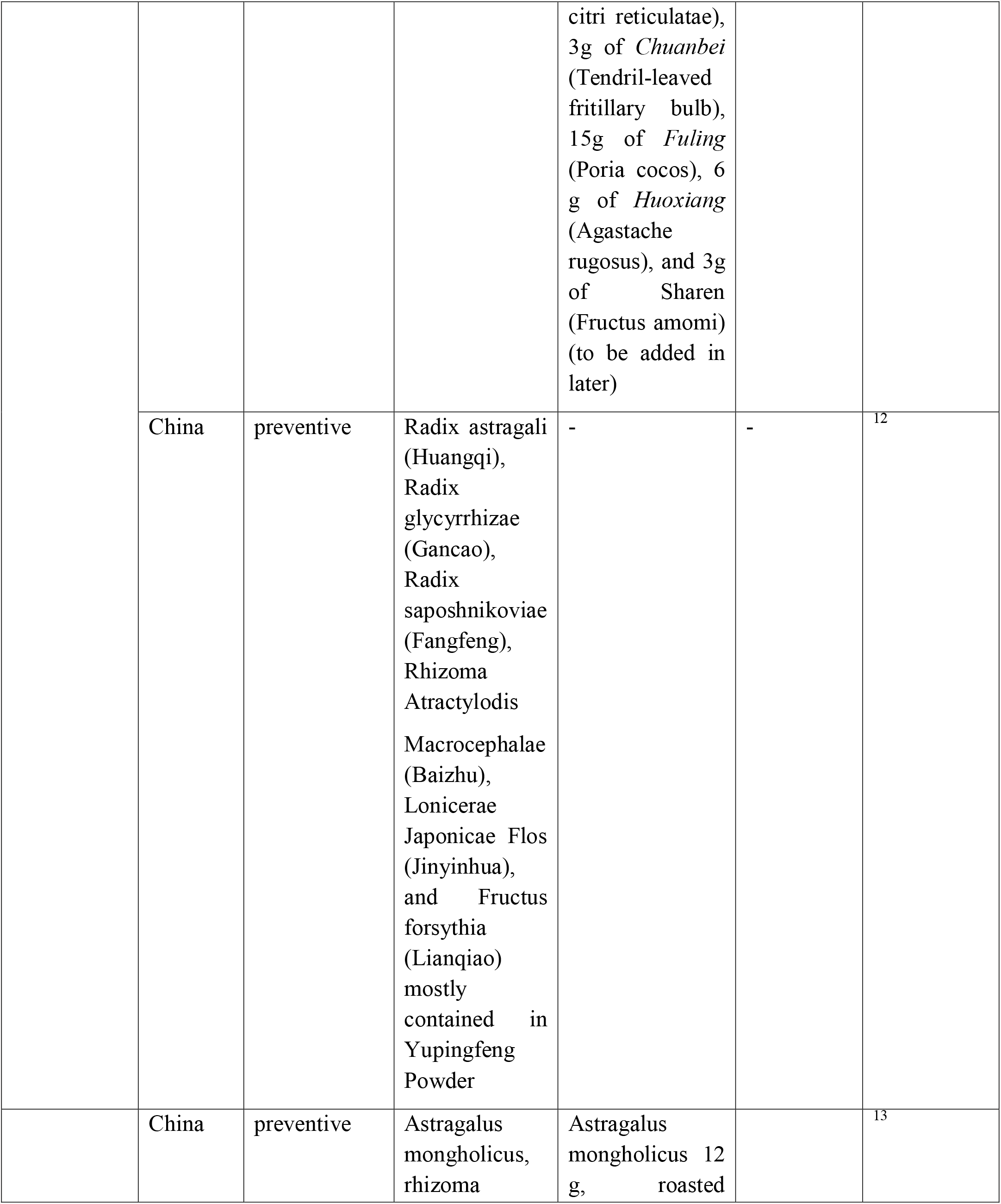
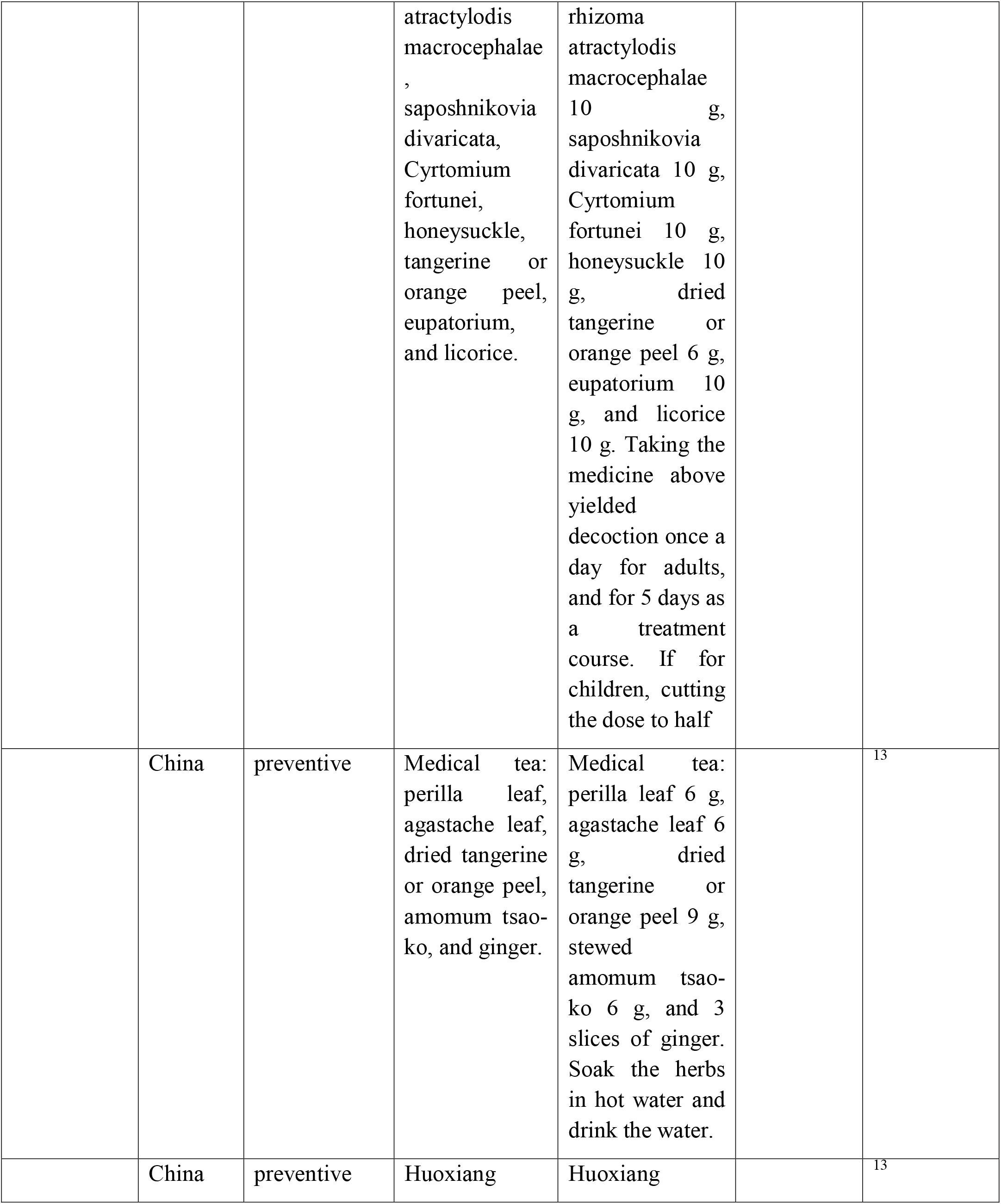
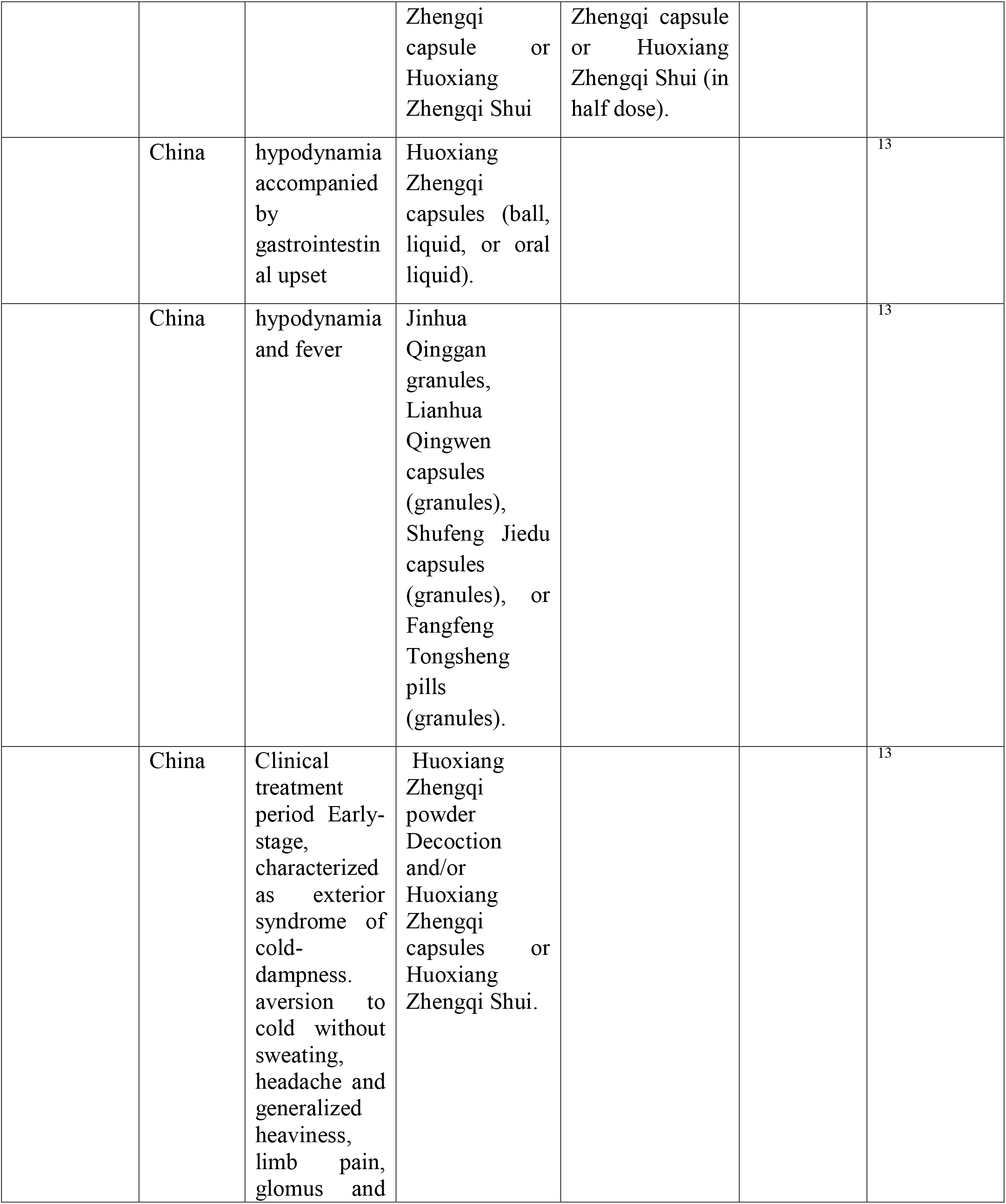
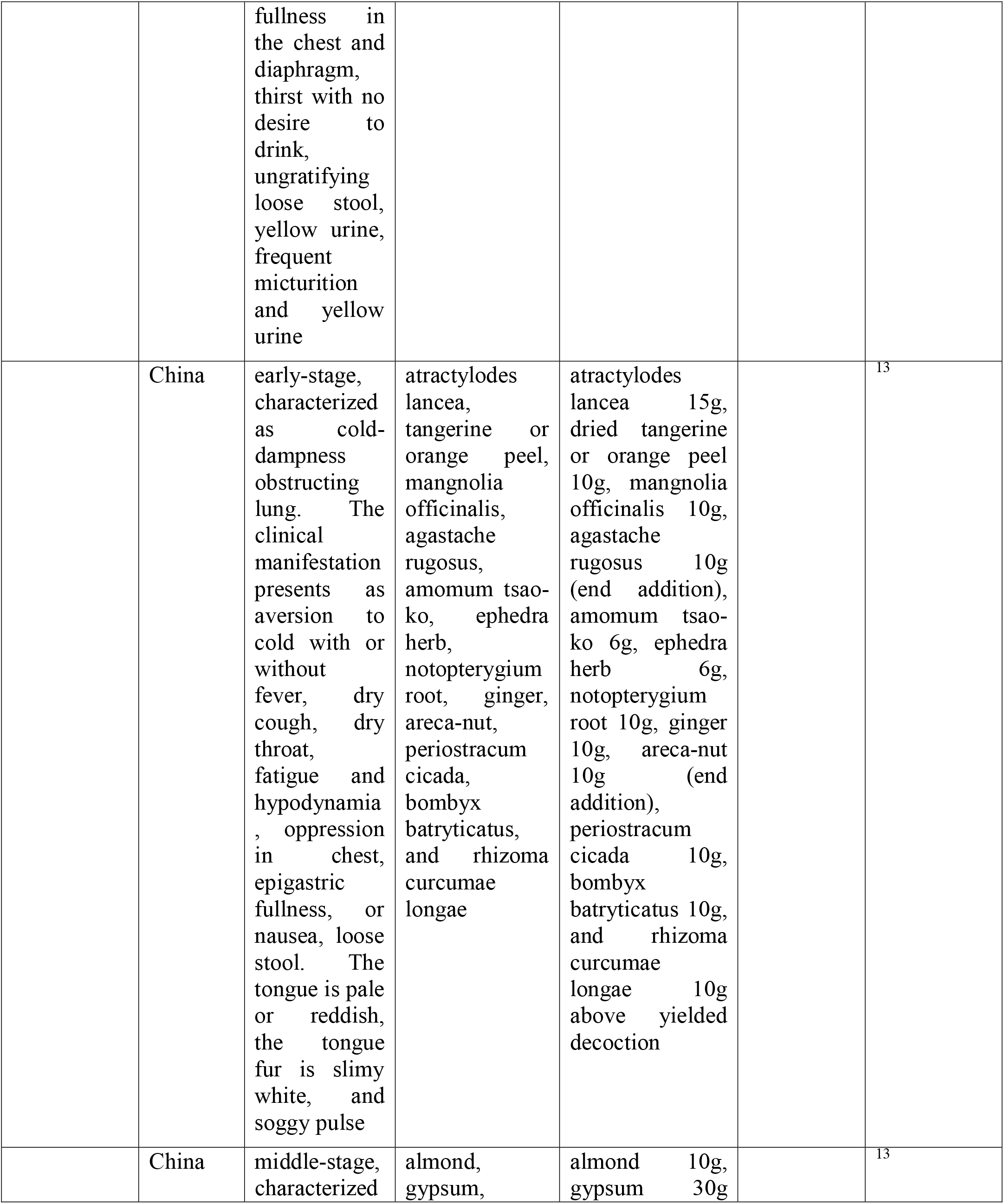
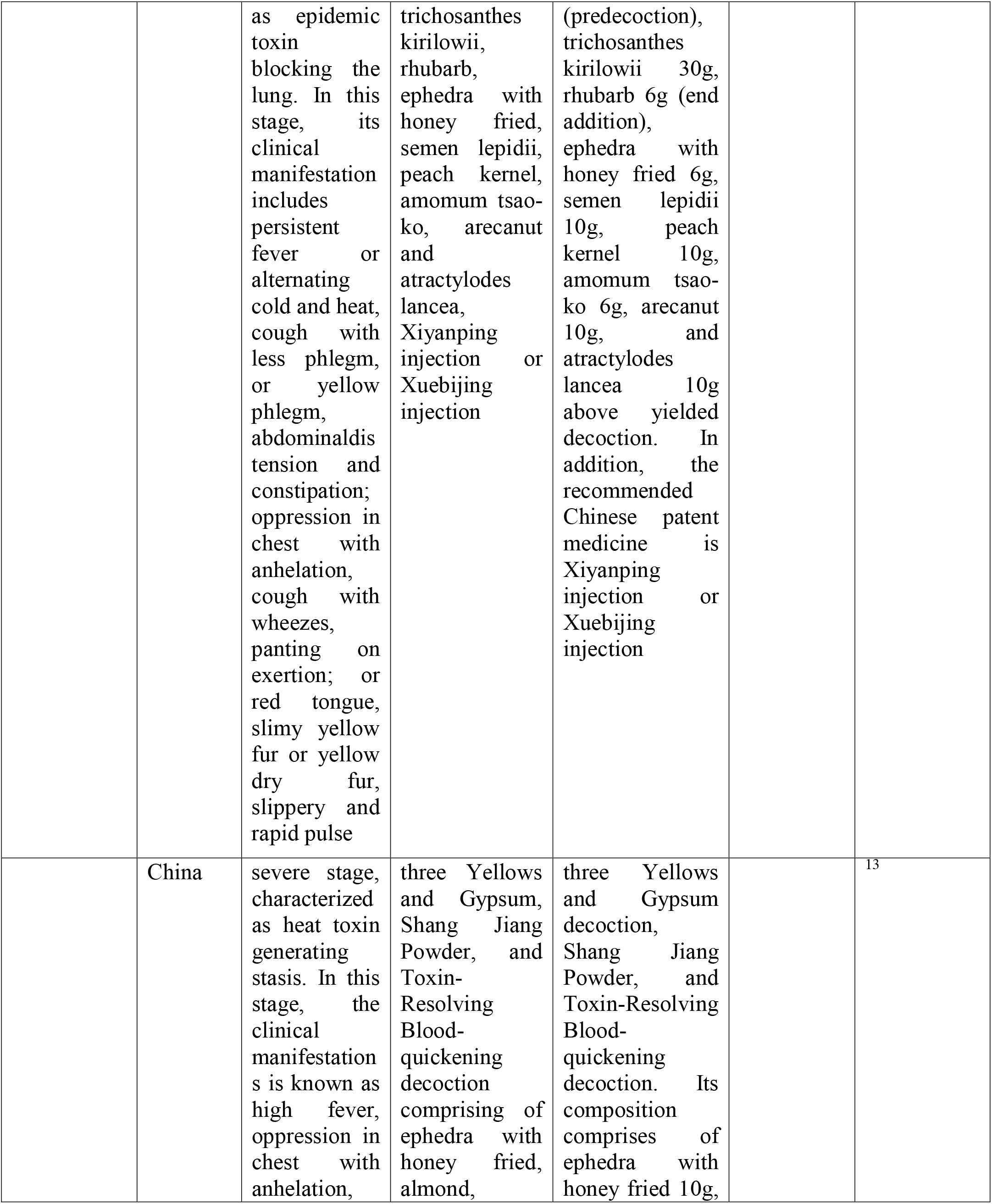
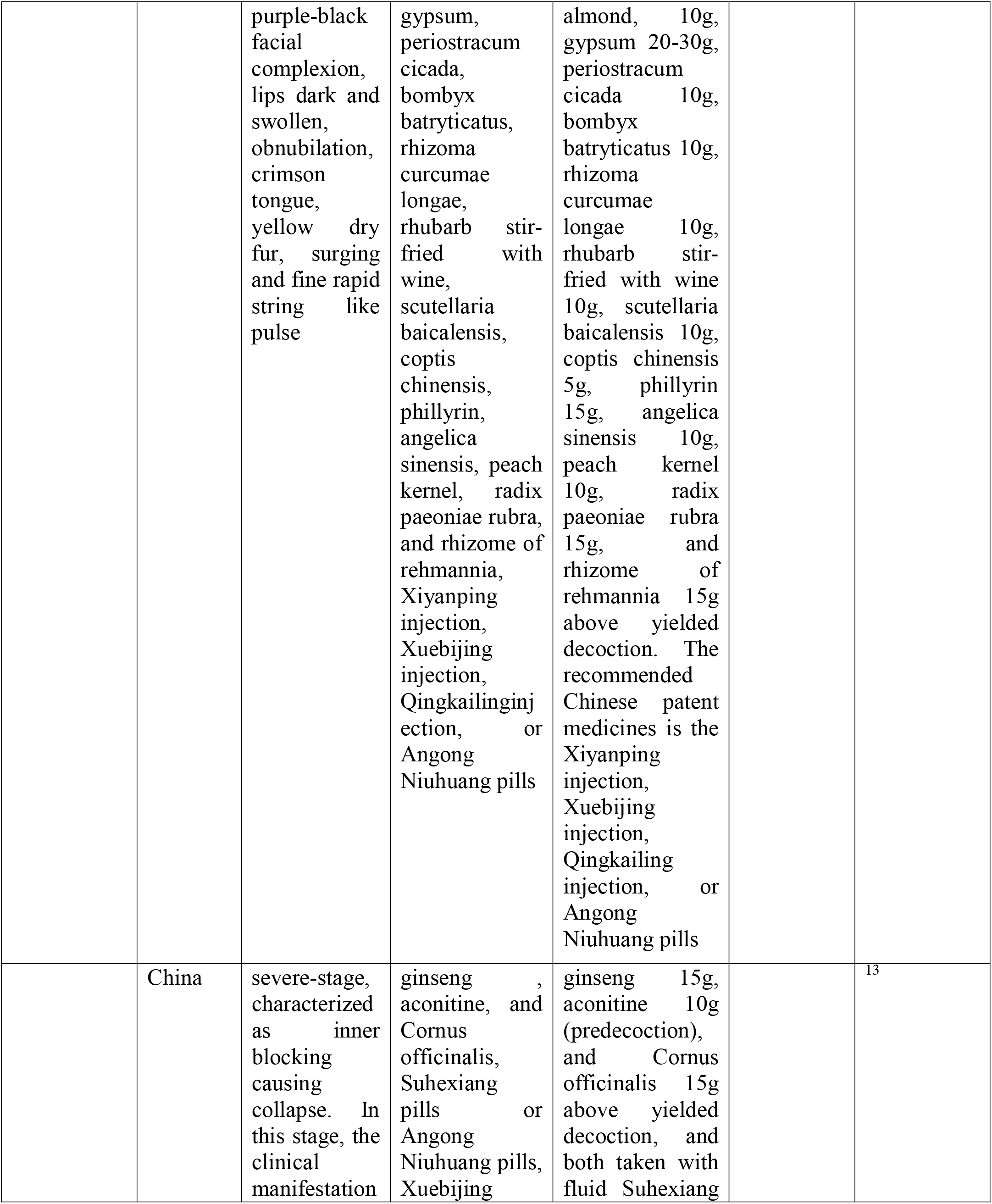
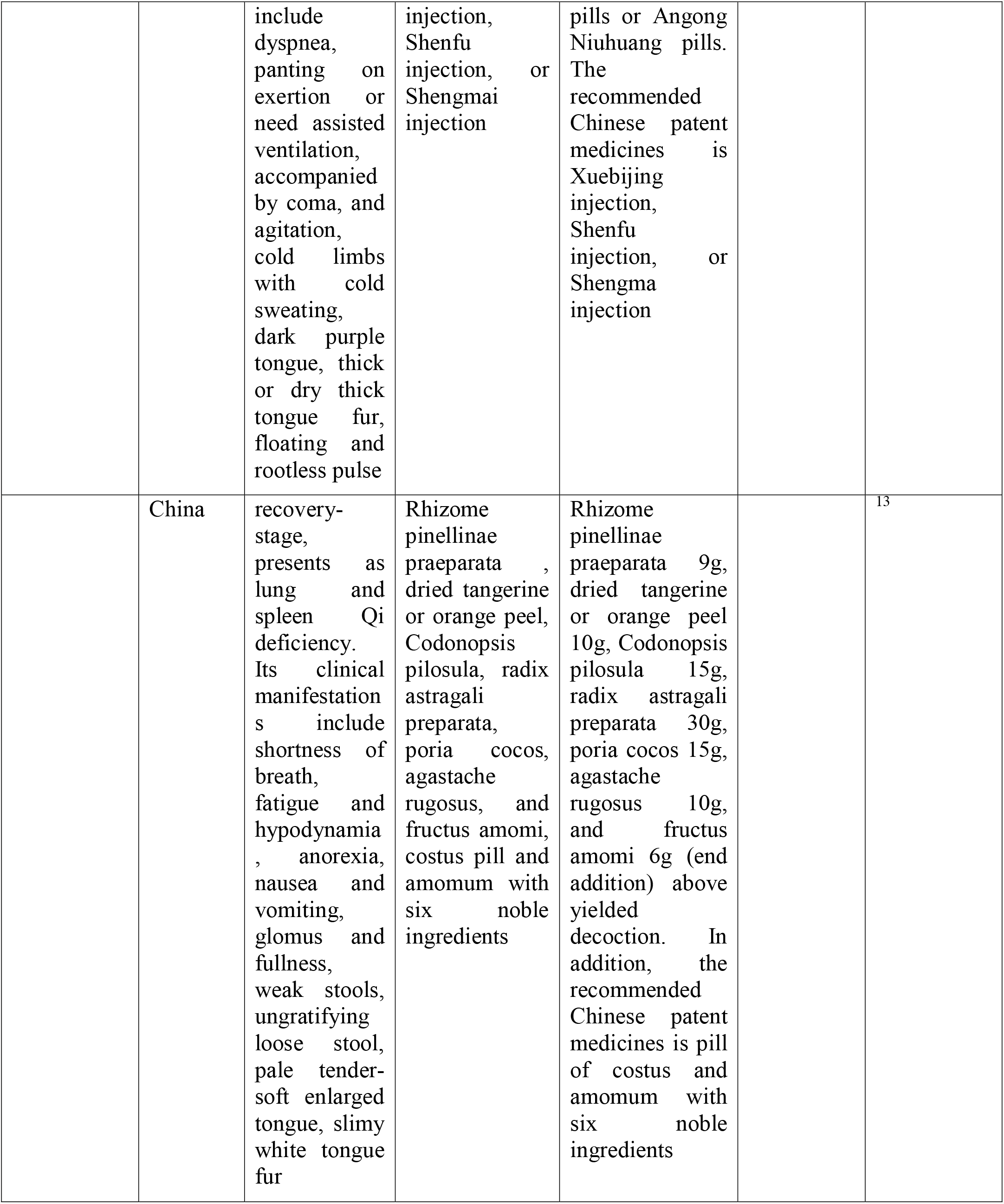
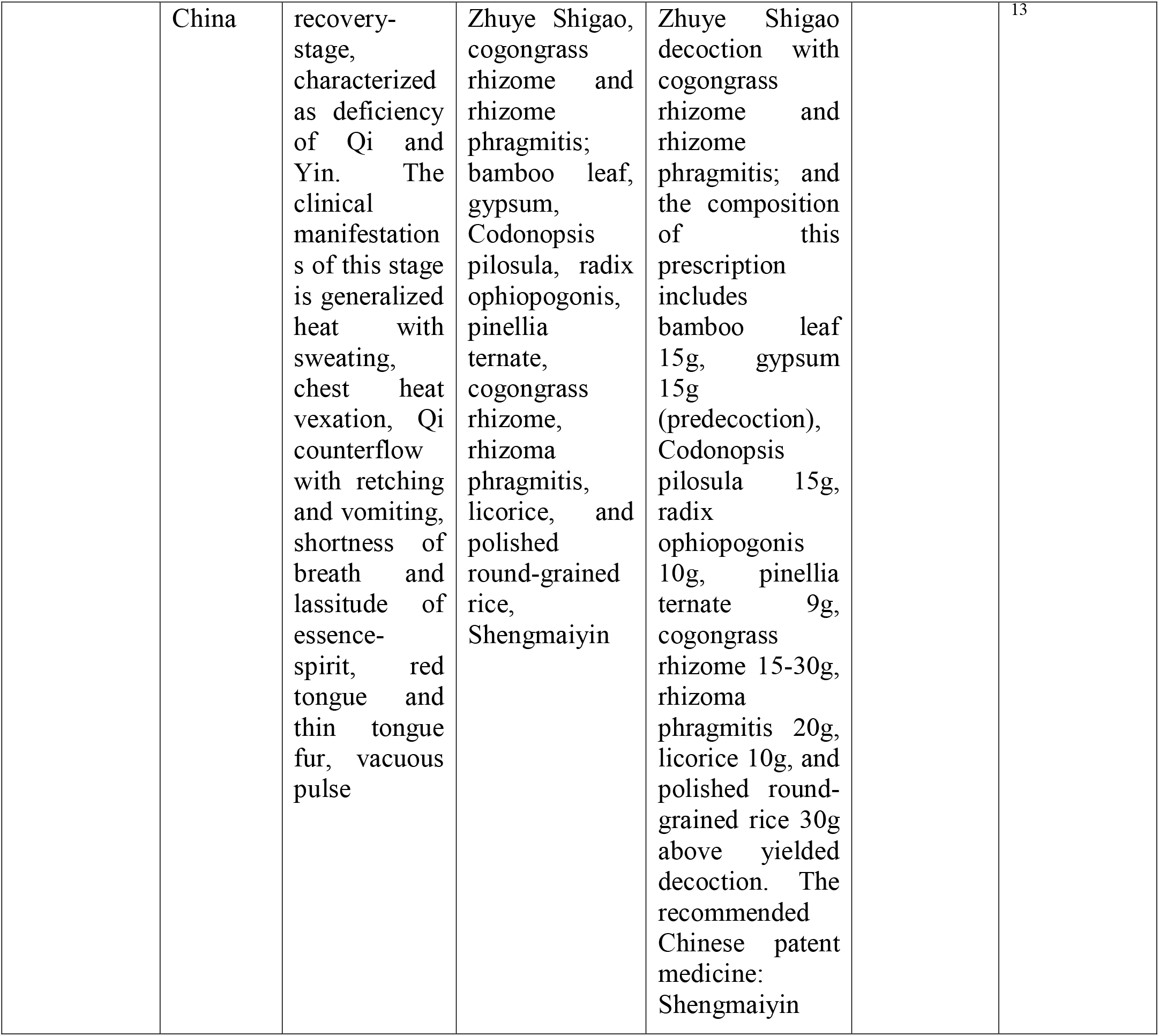
Describes the promotions country-wise, clinical treatment, medicines, drug dose and duration of the alternative therapy systems for COVID- 19.

## Ayurveda

### *Ark Ocimum sanctum* (*tulsi extract*)

It is said to prevent disease, promote general health, wellbeing and longevity. In addition to providing immunity, it is recommended as a treatment for a range of conditions including anxiety, fever, cough, asthma, diarrhea, vomiting, indigestion, dysentery, diabetes, arthritis, genitourinary disorders, and back pain.^14–16^

### Amritarishta

The main ingredient of Amritarishta is *Tinospora Cordifolia* and *Dashmool* with water. Dashmoolmatrabastiin has shown efficacy in the management of benign prostatic hyperplasia.^17^

### Agastya Harityaki

It contains Haritaki as one of the main components. The plant has been extensively used for cough, intermittent fevers, indigestion, anemia, asthma, neuropathy, etc. Haritaki has been well reported for its antiviral properties.^18^

### SamshamaniVati

It is an ayurvedic medicine used in the treatment of all types of fever. It is prepared with Giloya (*Tinospora cordifolia*).^19^ Giloya extract contains many constituents such as alkaloids, steroids, glycosides, and polysaccharides.^20^ Its extract is extensively used in various herbal preparations for the treatment of different ailments for its immunomodulatory, anti-inflammatory, anti-oxidant, anti-spasmodic, anti-osteoporotic, anti-arthritic, and anti-allergic properties.^21,22^

### *Trikatu* (*Pippali, Marich & Shunthi*) *powder and Tulsi leaves*

It is an herbal formulation containing dried fruits of *Piper nigrum* (Maricha) and *Piper longum* (Peepli), and dried rhizomes of *Zingiber officinale* (Sunthi) mixed together in equal quantities. Different extracts and fractions of Trikatu possess anti-oxidant,^23^ immunomodulatory,^24,25^ anti-allergic,^26^ and anti-arthritic^27^ activities. Studies report that Trikatu possesses the ability to improve the bioavailability of various drugs if combined with them thereby increasing the efficacy of the treatment. It is also prescribed for the treatment of asthma, cough, chronic rhinitis/ sinusitis etc.^28,29^

### *Ashwagandha* (*Withania somnifera*)

It is also called ‘Indian Ginseng’, is an important medicinal plant widely used against many diseases in Indian Systems of Medicine. It is shown to possess anti-microbial, anti-inflammatory, anti-tumor, anti-stress, neuroprotective, cardioprotective, and anti-diabetic properties.^30–32^ It has shown antiviral activity against infectious bursal disease virus and herpes simplex virus type 1.^33,34^

## Homeopathy

### Arsenicum album 30C

Arsenicum album 30C and two drops of sesame oil in each nostril each morning are suggested by the advisory given by Central Council for Research in Homeopathy (CCRH), India for prevention to COVID-19.^5^ Arsenicum album 30C is prepared by diluting aqueous arsenic trioxide until little or no arsenic remains, and is used in respiratory disorders. Mice intoxicated with arsenic trioxide injections when treated with Arsenicum album 30 showed revival and restoration of protein and DNA damage of liver and kidney.^35^ Some improvement in the clinical outcome of Acute Encephalitis Syndrome was observed with adjunctive Homeopathic medicines which included Arsenicum album 30.^36^

## Unani

### Sharbat Unnab

It is widely used in the Unani system of medicine as a blood purifier, curing sore throat, catarrh, cough, urticaria, chest pain and pneumonia.^37,38^

### Khamira Marwareed

It exhibits immune-stimulatory activity leading to a Th1-dominant immune state.^39^ Although considered to be cardio-protective no scientific evidence is reported yet.^40^

### Roghan Baboona

It exhibits anti-inflammatory and analgesic activities. Massage with Roghan Baboona is reported to give relief from symptoms and signs of osteoarthritis and rheumatoid arthritis in patients.^41^

### Arque-Ajeeb

It is a Unani formulation consisting of plant extracts of *Menthaarvensis* Linn, seeds extract of *Trachyspermumammi* Linn and Camphor. It is reputed for its beneficial effects in the treatment of diarrhea and cholera,^42^ but the claim of the efficacy is yet to be tested.

### Habb-e-Ikseer Bukhar

It is suggested as an antipyretic to lower the body temperature for clinical management of Dengue fever.^43^.

### Behidana

The entire plant is used for medicinal purposes, and it has been proven as anti-oxidant, anti-bacterial, anti-inflammatory, anti-allergic, aphrodisiac, nephroprotective, anti-atherosclerotic, anti-hypertensive, hypolipidaemic, hepatoprotective, anti-spasmodic, and anti-cancererous.^44^

### Sapistan

It exhibits significant anti-bacterial activity and is used to treat upper respiratory tract infections.^45^

### Darchini

It is also called Cinnamon, is a coagulant and prevents bleeding.^46^ It also possesses anti-microbial,^47^ anti-fungal,^48^ and anti-oxidan^49^ properties. Its use might be beneficial to reduce oxidative stress-induced complications and oxidative stress associated diseases.^50,51^

### Banafsha

It exhibits anti-inflammatory, anti-microbial, anti-oxidant, anti-pyretic, expectorant and anti-tussive, anti-spasm and bronchodilator, analgesic activities.^52^ Sharbatbanafsha also effectively relieves most of the complaints due to chronic sinusitis.^53^

### Chiraita

Previous research demonstrates that the *Swertia chirayita* extracts exhibit a wide range of biological activities, such as anti-bacterial, anti-fungal, anti-viral, anti-inflammatory, anti-diabetic and antioxidant.^54–56^

### Kasni

The root extract of Kasni (*Cichoriumintybus*) is shown to have anti-tumor and immunomodulatory properties^57^ Dried root is used as a tonic in fevers, rheumatic complaints vomiting, diarrhea, and enlarged spleen.^58^

### Afsanteen

It exhibits diverse pharmacological effects such as anti-helminthic, anti-bacterial, anti-pyretic and hepatoprotective.^59^

### Neem Bark

Its ingredients are used for the treatment of many cancers, infectious and metabolic diseases in traditional medicine.^60^

### SaadKoofi

SaadKoofi (*Cyperusrotundus*) is a medicinal plant having potential pharmacological actions both prophylactic and therapeutic.^61^

### *GuleSurkh* (Rosa damascene flower)

It is medicinally used in various diseases such as asthma, bronchitis, wounds, and ulcer.^62^ The mixture made by rose and honey is very effective in throat problems.^63^

## Traditional Chinese Medicine

Mostly combinations of TCMs were proposed for the prevention or treatment of COVID-19 which are already covered by several articles^4,5,8–10,12^ Since the therapeutic activity was not of an individual medicine but of cocktail they are not described here. However, early intervention of TCMs showed to be effective in improving cure rate of COVID-19 patients. It delayed the disease progression, shortened the course of the disease and reduced mortality in the treated patients.^4^ Based on the effective response of TCM on COVID-19 patients, TCM treatment strategies have been included in guidelines issued from China.^11,13^ Further details of TCMs used for the treatment of COVID-19 are described in Table 1.

## Discussion

The early cases of COVID-19 diseases were diagnosed in China in November 2019, which ultimately made a lockdown in a few regions of China in early 2020. Due to the unavailability of definite modern medicines for the prevention or treatment of COVID-19, China along with other methods of prevention or treatment promoted and treated 60,107 Chinese people with TCM. The effective cure rate of QPD against COVID-19 was reported ∼90%^4^. In India, the promotion of AMs (Ayurveda, Homeopathy, and Unani) is underway.^5^

The results of this study found that different medicines were proposed for the prevention or treatment of COVID-19 by different AM systems. All the proposals for promoting AMs were from China or India. TCMs were primarily promoted by China, whereas Ayurveda, Homeopathy, and Unani were promoted by India. Evidence from the screened records suggest that AMs were primarily considered as a preventive treatment of COVID-19 disease. The detailed mechanism of action of AMs on COVID-19 was not found in the studied records. We found that most of the medicines used were for the treatment of the symptoms (fever, cough, and chest related issues such as pneumonia, chest pain etc.) and boosting immunity. No RCT addressing the antiviral properties of these medicines against COVID-19 was observed. Robust evidence on the effect of these medicines on the COVID-19 life cycle and the infected individuals are lacking, and is required to understand their plausible role in the prevention or treatment of COVID-19.

## Conclusions

This study limits to selective AM options i.e. Ayurveda, Homeopathy, Unani, and TCM, however other uncovered AMs such as naturopathy, etc. needs to be explored. Scientific lab research and RCTs are lacking for the proposed AMs for COVID-19. Screening for potential candidates via *in silico* analysis using the bioinformatics tool followed by *in vitro* screening of the proposed medicines and RCTs are needed for scientific justification. A detailed investigation of the different combinations of these AMs is warranted to look if there is a definite solution to the COVID-19 disease. Studying other AMs for their efficacy to treat or prevent COVID-19, based on their known activities, would be an opportunity of window that could be explored. More information and research on AMs for COVID-19 treatment is needed from the different geographical locations of the World. This study may form an excellent foundation for developing lab research in the field of AMs and COVID-19, and draw the attention of scientists and researchers throughout the globe to study and understand the biological role of our listed AMs for the prevention and treatment of COVID-19. Following these suggestions, research conducted in this field may determine the importance and value of AMs and probable scope of a full systematic review.

## Data Availability

No external data is used.

## Acknowledgments

The authors are thankful to Dr. Nirmal Chandra Asthana for his continuous support and suggestions on the manuscript. The grammar was checked by Grammarly software and the plagiarism from the online plagiarism checker ‘Duplichecker’ (https://www.duplichecker.com/).

## Ethical Statement

None declared

## Authors’ contributions

AN reviewed and wrote the first draft of the review. VS conceived the study, reviewed and wrote the final version of the manuscript. ST reviewed the manuscript and gave critical suggestion.

## References

1. Xiao, Y. & Torok, M. E. Taking the right measures to control COVID-19. Lancet. Infect. Dis. (2020). doi:10.1016/S1473-3099(20)30152-3

2. Fauci, A. S., Lane, H. C. & Redfield, R. R. Covid-19 – Navigating the Uncharted. N. Engl. J. Med. (2020). doi:10.1056/NEJMe2002387

3. Coronavirus disease 2019 (COVID-19) Situation Report – 52.

4. Ren, J.-L., Zhang, A.-H. & Wang, X.-J. Traditional Chinese medicine for COVID-19 treatment. Pharmacol. Res. 155, 104743 (2020).

5. Advisory for Corona virus Homoeopathy for Prevention of Corona virus Infections Unani Medicines useful in symptomatic management of Corona Virus infection. (2020).

6. PRISMA TRANSPARENT REPORTING of SYSTEMATIC REVIEWS and META-ANALYSES.

7. Peters, M. D. J. et al. Guidance for conducting systematic scoping reviews. Int. J. Evid. Based. Healthc. 13, 141–146 (2015).

8. Stressed About Coronavirus? Here’s How Yoga Can Help. (2020).

9. National Health Commission of the People’s Republic of China.<Guideline on diagnosis and treatment of COVID-19 (Trial 6th edition).

10. Lu, H. Drug treatment options for the 2019-new coronavirus (2019-nCoV). Biosci. Trends (2020). doi:10.5582/bst.2020.01020

11. Shen, K. et al. Diagnosis, treatment, and prevention of 2019 novel coronavirus infection in children: experts’ consensus statement. World J. Pediatr. (2020). doi:10.1007/s12519-020-00343-7

12. Luo, H. et al. Can Chinese Medicine Be Used for Prevention of Corona Virus Disease 2019 (COVID-19)? A Review of Historical Classics, Research Evidence and Current Prevention Programs. Chin. J. Integr. Med. (2020). doi:10.1007/s11655-020-3192-6

13. Jin, Y.-H., Cai, L., Cheng, Z.-S., Cheng, H. & Deng, T. A rapid advice guideline for the diagnosis and treatment of 2019 novel coronavirus (2019-nCoV) infected pneumonia (standard version).

14. Jamshidi, N. & Cohen, M. M. The Clinical Efficacy and Safety of Tulsi in Humans: A Systematic Review of the Literature. Evid. Based. Complement. Alternat. Med. 2017, 9217567 (2017).

15. Singh, N. Tulsi: The Mother Medicine of Nature. International Institute of Herbal Medicine (2002).

16. Pattanayak, P., Behera, P., Das, D. & Panda, S. K. Ocimum sanctum Linn. A reservoir plant for therapeutic applications: An overview. Pharmacogn. Rev. 4, 95–105 (2010).

17. Gupta, A., Gupta, P. & Sharma, R. TO STUDY THE EFFICACY OF KANCHNAR GUGGULU AND DASHMOOL MATRA BASTI IN THE MANAGEMENT OF VATASHTHEELA W.S.R TO BENIGN PROSTATIC HYPERPLASIA. Int. J. Ayurveda Pharma Res. 7, 19–24 (2019).

18. Poudel, S. & Yadav, M. P. Agastya Haritaki Rasayana: A Critical Review. J. Drug Deliv. Ther. doi:https://doi.org/10.22270/jddt.v9i1-s.2283

19. Alsuhaibani, S. & Khan, M. A. Immune-Stimulatory and Therapeutic Activity of Tinospora cordifolia: Double-Edged Sword against Salmonellosis. J. Immunol. Res. 2017, 1787803 (2017).

20. Chemistry and medicinal properties of Tinosporacordifolia (Guduchi). Indian J. Pharmacol. 35, 83–91 (2003).

21. Panchabhai, T. S., Kulkarni, U. P. & Rege, N. N. Validation of therapeutic claims of Tinospora cordifolia: a review. Phytother. Res. 22, 425–41 (2008).

22. Sharma, U. et al. Immunomodulatory active compounds from Tinospora cordifolia. J. Ethnopharmacol. 141, 918–26 (2012).

23. Jain, N. & Mishra, R. Antioxidant activity of trikatu mega Ext. Int J Res Pharm Biomed Sci 2, 624–28 (2011).

24. Anthelmintic activity of water extracts of trikatu churna and its individual ingredients on Indian earthworms. Int J Pharm Bio Sci 3, 374–8 (2012).

25. Jain, N. & Mishra, R. Immunomodulator activity of trikatu. Int J Res Pharm Biomed Sci 2, 160–64 (2011).

26. Murunikkara, V. & Rasool, M. Trikatu, an herbal compound as immunomodulatory and anti-inflammatory agent in the treatment of rheumatoid arthritis--an experimental study. Cell. Immunol. 287, 62–8 (2014).

27. Maenthaisong, R. et al. Efficacy and safety of topical Trikatu preparation in, relieving mosquito bite reactions: a randomized controlled trial. Complement. Ther. Med. 22, 34–9 (2014).

28. Pattanaik, S., Hota, D., Prabhakar, S., Kharbanda, P. & Pandhi, P. Effect of piperine on the steady-state pharmacokinetics of phenytoin in patients with epilepsy. Phytother. Res. 20, 683–6 (2006).

29. Kasibhatta, R. & Naidu, M. U. R. Influence of piperine on the pharmacokinetics of nevirapine under fasting conditions: a randomised, crossover, placebo-controlled study. Drugs R. D. 8, 383–91 (2007).

30. Dar, N. J., Hamid, A. & Ahmad, M. Pharmacologic overview of Withania somnifera, the Indian Ginseng. Cell. Mol. Life Sci. 72, 4445–60 (2015).

31. Dutta, R., Khalil, R., Green, R., Mohapatra, S. S. & Mohapatra, S. Withania Somnifera (Ashwagandha) and Withaferin A: Potential in Integrative Oncology. Int. J. Mol. Sci. 20, (2019).

32. Grover, A., Agrawal, V., Shandilya, A., Bisaria, V. S. & Sundar, D. Non-nucleosidic inhibition of Herpes simplex virus DNA polymerase: mechanistic insights into the anti herpetic mode of action of herbal drug withaferin A. BMC Bioinformatics 12 Suppl 1, S22 (2011).

33. Pant, M & Ambwani Tanuj & Umapathi, V. Antiviral activity of Ashwagandha extract on Infectious Bursal Disease Virus Replication. Indian J. Sci. Technol. (2012).

34. Kambizi, L., Goosen, B.., Taylor, M. B. & Afolayan, A.. Anti-viral effects of aqueous extracts of Aloe ferox and Withania somnifera on herpes simplex virus type 1 in cell culture. S. Afr. j. sci 103, 359–360 (2007).

35. Kundu, S. N., Mitra, K. & Khuda Bukhsh, A. R. Efficacy of a potentized homeopathic drug (Arsenicum-Aalbum-30) in reducing cytotoxic effects produced by arsenic trioxide in mice: IV. Pathological changes, protein profiles, and content of DNA and RNA. Complement. Ther. Med. 8, 157–65 (2000).

36. Oberai, P. et al. Effectiveness of Homeopathic Medicines as Add-on to Institutional Management Protocol for Acute Encephalitis Syndrome in Children: An Open-Label Randomized Placebo-Controlled Trial. Homeopathy 107, 161–171 (2018).

37. Hs, K. & Amraz, I. New Delhi: CCRUM Dept of AYUSH Govt of India. (2005).

38. Talib, M., Aslam, M., Ahmed, M. A., Qamar, M. W. &, Shahid S. Chaudhary, A. J. Unnab: A boon to herbal nutraceuticals. Intrenational J. Adv. Aharmacy Medcine Bioallied Sci. (2017).

39. Khan, F., Ali, S., Ganie, B. A. & Rubab, I. Immunopotentiating effect of Khamira Marwarid, an herbo-mineral preparation. Methods Find. Exp. Clin. Pharmacol. 31, 513–22 (2009).

40. Ahmad, S. et al. Khamiras, a natural cardiac tonic: An overview. J. Pharm. Bioallied Sci. 2, 93–9 (2010).

41. Tanzeel, A. The Effect of Dalak (Massage) with Roghan-e-Baboona in Wajaul Mafasil (Arthritis) – A Clinical Study. A J. Siddha, Ayurveda, Unani, Yoga, Naturop. Homeopath. 4, (2004).

42. Khan, M. A., Khan, N. A., Qasmi, I. A., Ahmad, G. & Zafar, S. Protective effect of Arque-Ajeeb on acute experimental diarrhoea in rats. BMC Complement. Altern. Med. 4, 8 (2004).

43. Guidelines for Unani Practitioners for Clinical Management of Dengue Fever.

44. Jour, Fazeenah, H. & Quamri, M. BEHIDANA (CYDONIA OBLONGA MILLER.)-A REVIEW. World J. Pharm. Res. 5, 79–94

45. Latif, A., Tafseer, M. B., Rauf, A., Khan, A. U. & Rehman, S. LAOOQ SAPISTAN KHYAAR SHAMBARI-A UNANI HERBAL FORMULATION. Int. J. Pharm. Res. Bio-Science 2, 67–77 (2013).

46. Hossein, N., Zohrvand Zahra, M. A., Mahdi, S. & Ali, K. Effect of Cinnamon zeylanicum essence and distillate on the clotting time. J. Med. Plants Res. 7, 1339–1343 (2013).

47. Chang, S. T., Chen, P. F. & Chang, S. C. Antibacterial activity of leaf essential oils and their constituents from Cinnamomum osmophloeum. J. Ethnopharmacol. 77, 123–7 (2001).

48. Wang, S.-Y., Chen, P.-F. & Chang, S.-T. Antifungal activities of essential oils and their constituents from indigenous cinnamon (Cinnamomum osmophloeum) leaves against wood decay fungi. Bioresour. Technol. 96, 813–8 (2005).

49. Mancini-Filho, J., Van-Koiij, A., Mancini, D. A., Cozzolino, F. F. & Torres, R. P. Antioxidant activity of cinnamon (Cinnamomum Zeylanicum, Breyne) extracts. Boll. Chim. Farm. 137, 443–7 (1998).

50. Hussain, Z., Khan, J. A. & Rashid, H. Cinnamomum zeylanicum (Darchini): A Boon to Medical Science and a Possible Therapy for Stress-Induced Ailments. Crit. Rev. Eukaryot. Gene Expr. 29, 263–276 (2019).

51. Hussain, Z. et al. Protective effects of Cinnamomum zeylanicum L. (Darchini) in acetaminophen-induced oxidative stress, hepatotoxicity and nephrotoxicity in mouse model. Biomed. Pharmacother. 109, 2285–2292 (2019).

52. Qasemzadeh, M. J. et al. The Effect of Viola odorata Flower Syrup on the Cough of Children With Asthma: A Double-Blind, Randomized Controlled Trial. J. Evid. Based. Complementary Altern. Med. 20, 287–91 (2015).

53. Talat, H., Latafat, T., Aziz, M. N. & Yasmeen. A ranomized open comparative clinical study of sharbat ustukhuddus and sharbat banafsha in the management of chronic rhinosinusitis. Indian J. Tradit. Knowl. 19, 218–227 (2020).

54. Verma, H., Patil, P. R., Kolhapure, R. M. & Gopalkrishna, V. Antiviral activity of the Indian medicinal plant extract Swertia chirata against herpes simplex viruses: a study by in-vitro and molecular approach. Indian J. Med. Microbiol. 26, 322–6

55. Alam, K. D. et al. In vitro antimicrobial activities of different fractions of Swertia chirata ethanolic extract. Pakistan J. Biol. Sci. PJBS 12, 1334–7 (2009).

56. Chen, Y. et al. In vitro and in vivo antioxidant effects of the ethanolic extract of Swertia chirayita. J. Ethnopharmacol. 136, 309–15 (2011).

57. Hazra, B., Sarkar, R., Bhattacharyya, S. & Roy, P. Tumour inhibitory activity of chicory root extract against Ehrlich ascites carcinoma in mice. Fitoterapia 73, 730–3 (2002).

58. Chopra, R. N., Nayar, S. L. & Chopra, I. C. Glossary of Indian medicinal plants. (1982).

59. Khare, C. P. Encyclopaedia of Indian Medicinal Plants. (Springer Berlin, Heidenberg, New York, 2004).

60. Brahmachari, G. Neem--an omnipotent plant: a retrospection. Chembiochem 5, 408–21 (2004).

61. Hana, A. & Hifzul, K. Unani Perspective and New Researches of Sa’ad ku’fi (Cyperus rotundus): A Review. J. Drug Deliv. Ther. 8, 378–381 (2018).

62. Ghani, N. Khazainul Advia. Idara Kitabus Shifa, New Delhi. 1133–1135

63. R.N., C., I.C., C., K.L., H. & L.D, K. Indigenous drug of India. (HN Dhar and Sons pvt Limited. Calcutta, 1958).

64. Wang, Z., Chen, X., Lu, Y., Chen, F. & Zhang, W. Clinical characteristics and therapeutic procedure for four cases with 2019 novel coronavirus pneumonia receiving combined Chinese and Western medicine treatment. Biosci. Trends 14, 64–68 (2020).

